# Development and verification of non-supervised smartphone-based methods for assessing pure-tone thresholds and loudness perception

**DOI:** 10.1101/2024.06.25.24309468

**Authors:** Chen Xu, Lena Schell-Majoor, Birger Kollmeier

## Abstract

**Objective:** The benefit of using smartphones for hearing tests in a non-supervised, rapid, and contactless way has drawn a lot of interest, especially if supra-threshold measures are assessed that go beyond audiogram-based measures alone. It is unclear, nevertheless, how well these measures compare to more supervised and regulated manual audiometric assessments. The aim of this study is to validate such smartphone-based methods against standardized laboratory assessments.

**Design:** Pure-tone audiometry and categorical loudness scaling (CLS) were used. Three conditions with varying degrees of supervision were created and compared. In order to assess binaural and spectral loudness summation, both narrowband monaural and broadband binaural noise have been examined as CLS test stimuli.

**Study sample:** N = 21 individuals with normal hearing and N = 16 participants with mild-to-moderate hearing loss.

**Results:** The tests conducted here did not show any distinctions between smartphone-based and laboratory-based methods.

**Conclusions:** Non-supervised listening tests via smartphone may serve as a valid, reliable, and cost-effective approach, e.g., for pure-tone audiometry, CLS, and the evaluation of binaural and spectral loudness summation. In addition, the supra-threshold tests can be constructed to be invariant against missing calibration and external noise which makes them more robust for smartphone usage than audiogram measures.

## Introduction

Although the clinical routine audiometry tests (e.g., tone audiometry and speech audiometry) are highly valid and reliable to evaluate hearing ability, their practical drawbacks in terms of time consumption and personal intensiveness are not negligible (Colsman et al., 2020). Hence, employing a smartphone to conduct non-supervised listening tests - at least for simple routine cases where no medical supervision is required - might be a cost-effective alternative and attracts considerable interest. The current study aims at validating this approach by comparing non-supervised threshold and supra-threshold tests to classical laboratory-based audiometric assessments in a controlled way.

Previously, many studies have demonstrated that smartphone-based non-supervised methods are plausible and applicable to measure air-conduction pure-tone audiometry. Swanepoel et al. (2014) and Yousuf Hussein et al. (2016) developed and calibrated the hearScreen^TM^ app, examined in 15 normal hearing adults and 162 children, and compared it with clinical audiometry. Their results revealed that the smartphone- based pure-tone audiometry measurement was comparable to clinical audiometry. Later, Abu-Ghanem et al. (2016) evaluated the smartphone application ‘uHear’ for a questionnaire and a pure-tone audiometry test in the 26 participants aged 65 years and older and reported that there was an agreement between the app and audiometer assessment for most of the test participants in all frequencies. The app yielded a sensitivity of 100% and a specificity of 80% compared with clinical audiometry. More recently, Hazan et al. (2022) designed an experiment to test the reliability of a smartphone app ‘DuoTone’ on 1641 participants from a cloud database. Their results suggested that the test-retest reliability of the app did not differ from the standard audiometry performed in the clinics.

However, the validity and reliability of the smartphone tone audiometry apps are limited due to inherent limitations of the procedures employed. To the authors’ knowledge, nearly all of the current smartphone apps use either a modified Hughson-Westlake (Hughson et al., 1944) procedure or a not revealed procedure in order to circumvent any patent issues. The modified Hughson-Westlake procedure, most often employed in clinical audiometry, is widely adopted by clinicians due to its simple administration, little patient training, and easy implementation. Thus, most smartphone apps are directly adapted from clinical methods in order to be comparable with a clinical hearing test. However, according to the findings by Lecluyse and Meddis (2009) and Xu et al. (2023), the modified Hughson-Westlake procedure might be inaccurate and overestimate the true threshold if administered in a self-paced, unsupervised way due to occasional inattentiveness of the listeners. Following the recommendations of Lecluyse and Meddis (2009) and Xu et al. (2023), the present study therefore adopts the non-clinical adaptive procedure (e.g., the single interval up and down procedure SIUD, proposed by Lecluyse & Meddis, 2009) to assess air-conduction pure-tone audiometry on a smartphone and compares the acquired results with the laboratory-based measurements.

Another limitation of smartphone apps to measure the individual audiogram is their dependence on an absolute calibration of the earphones employed which can not be warranted, e.g., for Android devices. This problem is mostly circumvented by using supra-threshold tests (e.g., digit-in-noise (DIN) test, Smits et al., 2004) that assess a certain relative quantity of stimulus components (e.g., signal-to-noise ratio at threshold) and are largely independent of the absolute presentation level. Hence, supra-threshold auditory measures (e.g., speech-in-noise (SIN) tests) have attracted much attention in the last years for hearing screening via smartphone.

In clinical audiology, such supra-threshold tests are used to specify individual functional deficits. Of these, the assessment of loudness growth with increasing level or stimulus bandwidth is of clinical interest, e.g., determining the recruitment phenomenon and for fitting hearing devices (Kollmeier & Hohmann, 1995; Oetting et al., 2016; Koppun et al., 2022). Individual loudness perception is commonly measured employing the categorical loudness scaling (CLS) technique and quantified with a monotonic loudness growth function (Brand & Hohmann, 2002; Oetting et al., 2014). The task of the CLS requires participants to select the descriptors from an 11-point scale, e.g., ‘too loud’, ‘medium’, ‘soft’, etc., based on their loudness perception. The CLS is a supra- threshold listening test that has been included in the ‘auditory profile’ (i.e., a comprehensive and well-specified set of audiological test procedures described in Van Esch et al., 2013) and has also recently been proposed for usage in machine-learning-supported auditory profiles by Saak et al. (2022).

The standardized adaptive procedure to perform CLS measurement (i.e., Adaptive Categorical Loudness Scaling, **ACALOS**) was introduced by Brand and Hohmann (2002) and standardized in ISO 16832 (2006). CLS has a broad application in clinical audiology, not only as a diagnostic tool but also to fit hearing aids or cochlea implants. For diagnostic purposes, an increase in loudness growth with stimulus level – clinically termed as recruitment phenomenon and assumed to be due to dysfunctional outer hair cells (Hallpike & Hood, 1959; Buus & Florentine, 2002) – can well be characterized by CLS (e.g., Kollmeier & Hohmann, 1995, Launer, 1995, and Rasetshwane et al., 2015). Jürgens et al. (2011) proposed to estimate the hearing loss attributable to outer hair cells (OHC) by applying CLS and concluded that CLS could be a measure of auditory nonlinearity. Further diagnostical applications of CLS were described, e.g., by Shiraki et al. (2022) as a means to better characterize patients with certain patterns in Bekesy audiometry and by Erinc et al. (2022) and Hébert et al. (2013) as a means to better characterize patients with tinnitus and hyperacusis.

With respect to using CLS as a tool for hearing device fitting, many studies have demonstrated that individualized loudness compensation for narrowband signals can lead to a better-individualized treatment with hearing devices (see Kollmeier & Hohmann, 1995, Kollmeier & Kießling, 2018, Oetting et al., 2018, and Fereczkowski et al., 2023 for hearing aids and Müller-Deile et al., 2021 for cochlea implants). Despite its theoretical advantage to characterize supra-threshold functional hearing deficits and of the compensation by an appropriately, individually fitted hearing device, the usage of CLS for clinical purposes has been limited due to several reasons:

a. Time constraints in clinical settings that interfere with the usage of more sophisticated methods beyond the minimum set of clinical routine procedures. However, self-paced, smartphone-based procedures might take over that do not impose such a time-consuming burden on the professional audiologists.
b. Previous forms of CLS have been discredited by an influential paper by Elberling (1999) arguing that the uncertainty in hearing aid gain setting will not be reduced by CLS. However, their claim was based on the questionable assumption of a perfectly-known individual threshold. More refined measuring and evaluation techniques in CLS (e.g., Brand & Hohmann, 2002, and Oetting et al., 2014, 2016) demonstrate a low correlation between scaling slope estimate and individual threshold, thus demonstrating the importance of the individually obtained loudness growth function for hearing loss compensation.
c. Recent insights into the individually strongly varying loudness summation across frequency and across ears as demonstrated by Oetting et al. (2016) who reported that using narrowband gain compensation, levels of HI listeners to reach ‘medium loud’ were lower than for NH listeners when broadband signals were presented. Participants with the same hearing thresholds perceived loudness substantially differently for binaural broadband signals. Thus Oetting et al. (2016) recommended that the broadband and binaural loudness scaling should be included for hearing-aid fitting. To further investigate the potential consequences of the spectral and binaural loudness summation, Van Beurden et al. (2018) extended the study of Oetting et al. (2016) by recruiting more test participants with a broader range of hearing loss. Spectral loudness summation of HI listeners was detected to be greater than of NH listeners for both monaurally and binaurally presented signals. The effect of hearing loss did not significantly influence the binaural loudness summation. In agreement with Oetting et al. (2016), Van Beurden et al. (2018) found large individual variations in HI listeners for binaural broadband signals. In this study, we, therefore, follow the recommendations by Oetting et al. (2016) and Van Beurden et al. (2018, 2021) to employ not only narrowband signals presented unilaterally, but also broadband signals presented bilaterally for both NH and HI listeners.

Even though CLS is an applicable and useful measurement for clinical diagnostics and assessment of hearing loss compensation as introduced above (e.g., Rasetshwane et al., 2015; Fultz et al., 2020), it is not yet accessible for a smartphone or any other mobile device. There is only one study published so far that introduced a remote CLS measurement on a laptop and compared it with the laboratory setting (Kopun et al., 2022). However, they did not examine the test persons via smartphone and did not consider HI participants. Furthermore, Kopun et al. (2022) only measured 5 participants for the validation study. One possible obstacle against self-controlled CLS measurement in an unrestricted environment is the influence of background noise (which might cause a bias at low stimulus levels that might be confused with a recruitment phenomenon) or any inattention effect of the participant (as simulated in Xu et al., 2023). Hence, in this paper, one of our objectives is to examine the plausibility and validity of the smartphone-based app for CLS measurement under different degrees of control in experimental settings.

Taken together, the following research questions should be answered by our study by performing three sub-experiments (i.e., Exp 1: pure-tone audiometry; Exp 2: adaptive categorical loudness scaling; Exp 3: binaural and spectral loudness summation) that all employ normal-hearing and hearing-impaired listeners and compare laboratory situations with self-steered, smartphone-based setups:

- Are the results of the smartphone-based pure-tone audiometry and categorical loudness scaling quantitatively comparable to a laboratory-based assessment when using various statistical measures (e.g., correlation coefficient R, root mean square error, etc.)?
- Which factors (e.g., the way of supervision, the degree of hearing loss, and test frequency) might influence the differences between smartphone-based and laboratory- based measurements?
- Is the smartphone test able to detect individual differences in binaural and spectral loudness summation in a similar way as laboratory-based measures?

## Materials and methods

### Pure-tone audiometry

Pure-tone audiometry was assessed via a single-interval-up-and-down (SIUD) procedure, introduced by Lecluyse and Meddis (2009). Listeners were presented with a probe tone and a cue tone which had a 10 dB higher sound level than the probe tone and had a 20% chance to be muted, and required to indicate how many tones they have heard. The smartphone user interface of SIUD is provided in the left bottom corner of Fig. 1a. If listeners answered correctly, the task became harder by decreasing the sound level of the following trial. In the end, the track converged at the level of the listener’s hearing threshold. The behavioral data were fitted to a logistics psychometric function and the level (L_50_) corresponding to 50% of the psychometric function was estimated as the hearing threshold.

**Fig. 1.**
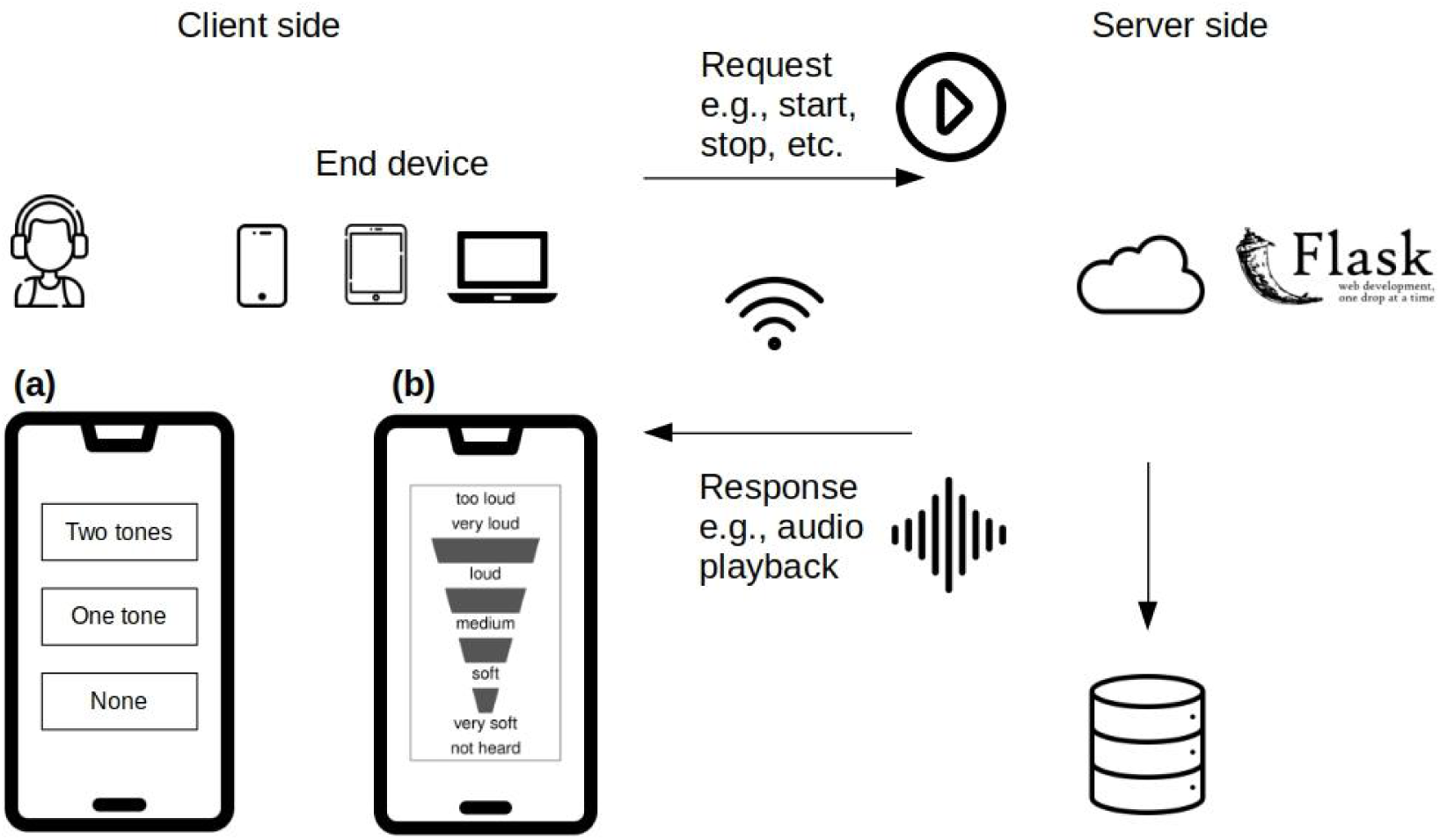
Overview of the smartphone application. On the client side, the user interface of two assessments (i.e., (a) pure-tone audiometry and (b) categorical loudness scaling) is shown. On the server side, the web application framework ‘FLASK’ is available for processing requests from a listener. The measurement data was not stored locally but in the cloud database.

The stimuli were pure tones consisting of 0.2 s duration for each tone, 20 ms cosine ramps, and 0.2 s for a break between two tones at 0.25, 1, and 4 kHz frequencies for both ears. The starting level for the probe tone was set up at 50 dB with a random offset between 0 and 5 dB. There was a fixed level difference of 10 dB between the probe and cue tone. If listeners reported that the first stimulus was not heard, the initial level increased until it was audible. The initial step size was chosen as 10 dB and reduced to 2 dB after the first reversal. The track terminated when at least 14 reversals and 10 trials were reached. The trials before the 4th reversal were excluded from the threshold calculation.

### Adaptive categorical loudness scaling

Adaptive categorical loudness scaling (ACALOS), described by Brand and Hohmann (2002) and ISO 16832 (2006), was applied to measure an individual’s loudness growth function. There were in total 11 categorical scale values distributed on the 50-point categorical units (CU) scale according to Heller (1985), i.e., the verbal values ‘very soft’ (5 CU), ‘soft’ (15 CU), ‘medium’ (25 CU), ‘loud’ (35 CU) and ‘very loud’ (45 CU), four intermediate categories without verbal labels, and the two limiting categories ‘not heard’ (0 CU) and ‘too loud’ (50 CU) in ACALOS. Listeners needed to rate the stimuli based on their individual loudness perception given the 11 categories. The user interface for the smartphone of ACALOS is shown in the left bottom corner of Fig. 1b. ACALOS mainly comprised two phases, i.e., ‘dynamic range estimation’ and ‘presenting and re-estimation’. During dynamic range estimation, the procedure started at 65 dB and presented upward and downward stimuli in an interleaved manner to obtain a rough estimate of the dynamic range between 0 CU and 50 CU. The individual loudness function was then fine-tuned in the second phase by presenting stimuli at 5 levels estimated from the first phase corresponding to the categorical loudness of 5, 15, 25, 35, and 45 CU in a randomized order.

The ‘BTUX’ method, which was introduced by Oetting et al. (2014), was employed to fit a loudness growth function. The descriptive parameters (i.e., hearing threshold level (HTL) corresponding to 2.5 CU, median loudness level (MLL at 25 CU), and uncomfortable loudness level (UCL at 50 CU), respectively) were derived from the fitted loudness growth function (Oetting et al., 2014). Furthermore, the most comfortable loudness (MCL) was estimated as the sound level at 20 CU of the growth function (Van Esch et al., 2013). Finally, the dynamic range (DR) was calculated as the difference between UCL and HTL.

The narrowband stimuli were one-third-octave-band low-noise noises (Kohlrausch et al., 1997) centered at 0.25, 1, and 4 kHz (later referred to as LNN250, LNN1000, and LNN4000, respectively). The broadband stimulus was uniformly exciting noise (UEN17) with equal energy in each of the 17 critical frequency bands, defined in Zwicker (1961). All stimuli (i.e., three narrowband and one broadband stimuli) were presented monaurally for both ears. In addition, LNN1000 and UEN17 were played bilaterally. The duration for all signals was 1 s with 50 ms rise and fall ramps.

### Smartphone application design

Fig. 1 shows the overview of the employed smartphone application. The web-app was developed based on the flask (version 1.1.2) framework in python (Python Software Foundation, version 3.10.6) while the database was based on SQLite3 (version 3.37.2). Both frameworks are open source.

The workflow to conduct a non-supervised listening test on a smartphone was as follows: the listener first registered an account and signed up to the dashboard. There are some general instructions, e.g., study background, user consent, and test environments, displayed in text format on the dashboard. Then the listener needed to indicate which measurement to perform by clicking on the appropriate button.

Subsequently, some specific guidelines for the chosen listening test were shown. The listener started the measurement by clicking on the ‘start’ button and the stimuli were automatically presented to the listener. The listener considered and later responded to questions, i.e., ‘How loud was the sound?’ or ‘How many tones have you heard?’ for CLS and pure-tone audiometry assessments, respectively. The response data were returned to the server via WLAN and stored in the cloud database. Based on the incoming response, the server prepared the adjusted stimulus (here, the adjustment mainly refers to the sound levels for both listening tests) and played it back to the listener. The listener was redirected to the dashboard when the listening test was completed. No data were stored locally on the smartphones but, instead, were primarily stored on the server.

### Subject groups

21 normal hearing (NH, aged between 20 and 35 years; 7 males, 14 females) and 16 hearing impaired listeners (HI, aged between 67 and 88 years; 11 males, 5 females) participated in the study. The participants in the NH group are mainly members of the working group and students of the university. The HI listeners were recruited via the database of Hörzentrum Oldenburg gGmbH. The mild-to-moderately impaired listeners with sensorineural hearing loss exhibited pure-tone averages (PTA) varying between

26.3 and 42.5 dB while NH listeners yielded thresholds at or below 15 dB for all frequencies between 250 Hz and 4 kHz. The differences in PTA between the left and right ears of HI listeners did not exceed 10 dB, indicating that the hearing loss of all HI listeners was symmetric. All participants did not have any previous experience with smartphone hearing tests. The listeners received an expenditure compensation of 12 euros per hour for their participation in the study. The research ethics committee of the University of Oldenburg approved the proposal (Drs. EK/2022/011) for this study.

Table 1. provides the means and standard deviations of the clinical audiogram (IEC 60645-1, 2002) as a function of 11 frequencies, together with pure tone average (PTA), for both ears of HI listeners measured by an audiologist with HDA200 headphones. The PTAs for better ears of HI listeners were 31.8 (± 5.3) while the mean PTA difference was less than 2 dB.

**Table 1.** Summarized statistics (i.e., average and standard deviation) on pure-tone thresholds (in dB HL) of the group of hearing-impaired listeners (N =16) for both ears with frequency varying from 0.125 to 8 kHz (11 frequencies) measured by the clinical audiogram (IEC 60645-1, 2002).

|  |  | Frequency (Hz) |  |  |  |  |  |  |  |  |  |  |  |
| --- | --- | --- | --- | --- | --- | --- | --- | --- | --- | --- | --- | --- | --- |
|  |  | 125 | 250 | 500 | 750 | 1000 | 1500 | 2000 | 3000 | 4000 | 6000 | 8000 | PTA<sup>a</sup> |
| Left | Ave. | 12.5 | 12.1 | 18.9 | 22.9 | 24.6 | 31.4 | 38.9 | 49.6 | 53.9 | 59.6 | 67.1 | 34.1 |
|  | SD | 7.3 | 7.8 | 7.9 | 8.9 | 7.5 | 7.9 | 8.4 | 9.5 | 10.0 | 8.0 | 7.3 | 4.9 |
| Right | Ave. | 15.0 | 16.1 | 21.4 | 23.6 | 24.3 | 30.7 | 35.3 | 45.0 | 50.0 | 56.4 | 66.1 | 32.8 |
|  | SD | 9.0 | 10.6 | 9.3 | 9.1 | 8.1 | 9.2 | 9.1 | 10.7 | 10.0 | 9.1 | 9.2 | 5.5 |

<sup>a</sup> PTA (i.e., pure-tone average) denotes the average thresholds of 0.5, 1, 2, and 4 kHz

### Test conditions

Table 2. reports the difference in the experimental design of the three conditions. Overall, the experimental design is a repeated measures design. The main difference among the different conditions was the degree of supervision. Condition I was a fully-supervised, manual measurement as reference. Condition III was a non-supervised assessment. Condition II was semi-supervised, i.e., the test examiner was available on request for questions while the experiment ran automatically under the control of the same adaptive procedure as for condition III. Specifically, the examiner did not have access to the log data in condition II and only answered questions.

**Table 2.**
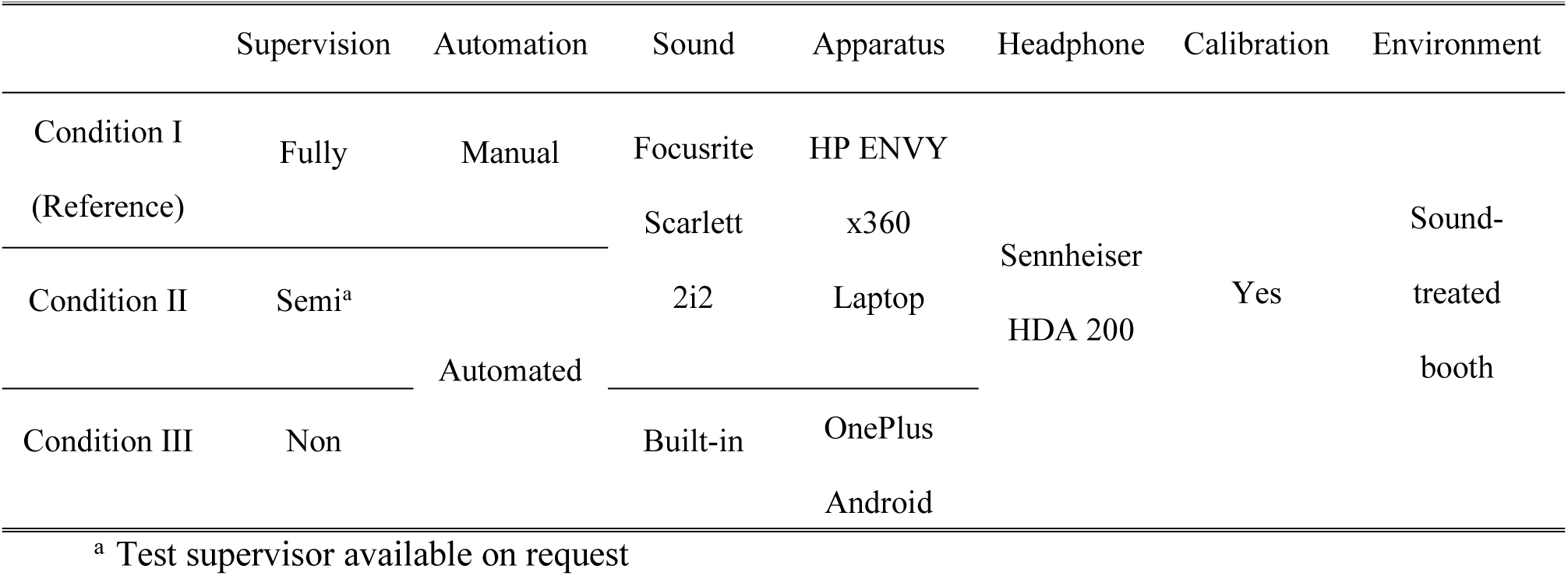
Experimental design for the three conditions employed that differed in the degree of supervision. Condition I implements a fully-supervised, manual laboratory measurement while condition III implements a non-supervised, automatic smartphone assessment, and condition II (“in-between condition”) represents a semi-supervised condition using a self-controlled data acquisition on a laboratory setup. All three experiments had the same acoustic environment (i.e., a sound-treated listening booth).

Furthermore, the sound card for conditions I & II was Scarlett 2i2, while the built-in sound card of the smartphone was employed in condition III. HP ENVY x360 laptop was used for conditions I & II while the Android smartphone (OnePlus Nord N10 5G 128 GB, google chrome downloaded) was used for condition III. The same calibrated smartphone was provided to all participants.

Finally, in all three conditions the same HDA200 headphone was employed in a sound-attenuated booth. All conditions were calibrated employing a B&K artificial ear 4153, a B&K 0.5-inch microphone 4134, a B&K microphone pre-amplifier 2669, and a B&K measuring amplifier 2610. The target level for calibration was 80 dB SPL.

### Data analysis

#### Psychophysical parameters

As already mentioned before, L_50_ (i.e., the half-way point of the psychometric function) in the pure-tone audiometry experiment was estimated as the hearing threshold, described in Eq. 1:

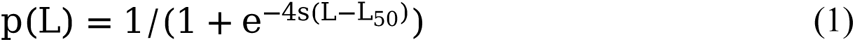

where p(L) is the probability of correct responses, L defines the sound level, and s denotes the slope of the half-way point of the function. Moreover, the signed difference between condition II or III, respectively and I is defined as L_50,II/III_ − L_50,I_, where L_50,II/III_ denotes the hearing threshold measured in condition II or III, respectively, while L_50,I_ is the hearing threshold measured in condition I. In addition, the absolute value of the difference is described as L_50,II/III_ − L_50,I_ . Finally, the root mean square error (RMSE) and the Pearson correlation coefficient (R) of conditions II and III against I are calculated.

For the categorical loudness scaling experiment, loudness functions as defined in Brand and Hohmann (2002) and Oetting et al., (2014) were employed (cf. Eq. 2), which consist of two linear parts and one transition region using a Bezier fit:

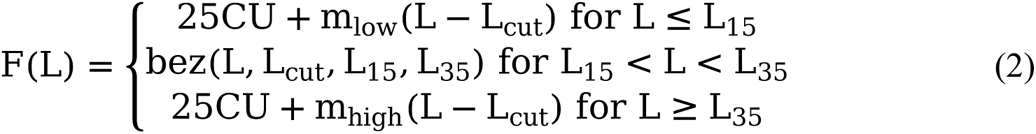

where m_low_ and m_high_ denote the slope value of the low and high linear part, L_cut_ is the intersection level of the two linear parts, L_15_ and L_35_ are the levels of the ‘soft’ and ‘loud’ category respectively, and bez is a quadratic smoothing function between L_15_ and L_35_. The Pearson correlation coefficient (R), root mean square error (RMSE), and bias of levels for each category (in total 11 categories) are calculated. For binaural loudness summation, the level difference for equal loudness (LDEL) is calculated as:

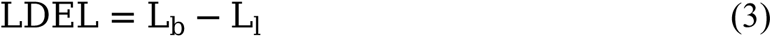

where L_b_ and L_l_ are defined as the level for binaural and monaural presentation of the **left ear** at the same category unit (i.e., equal loudness) respectively. The LDEL of the **left ear** for spectral loudness summation is described as:

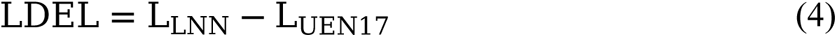

where L_LNN_ and L_UEN17_ denote the level for low-noise narrowband noise and UEN17 broadband noise at the same category unit respectively. All algorithms for experimental data fitting were developed in MATLAB R2021a (The MathWorks, Inc., Natick, MA).

#### Statistical analysis

A mixed-design ANOVA was applied using degree of hearing loss (two levels: NH/HI) as a between-subject factor, condition (three levels: I/II/III), and frequency (three levels: 0.25, 1, and 4 kHz) as within-subject factors. Furthermore, a post-hoc analysis among conditions using a pair-wise t-test was carried out, where the p value was corrected with ‘Bonferroni’. In the post-hoc analysis, condition I was set up as a reference group. If p value < 0.05 (*), 0.01 (**), 0.001 (***), and 0.0001 (****), the statistical test is considered as being significant, highly significant, very highly significant, and extremely significant, respectively, while if p value >= 0.05 (ns), the result is not significant, implying that there is no difference between two conditions. The ‘Tidyverse’ package (Wickham et al., 2019) developed in the software environment ‘R’ (R Foundation for Statistical Computing) was employed for the statistical analysis of the mixed-design ANOVA and the post-hoc analysis.

## Results

### Experiment I: Pure-Tone Audiometry

Fig. 2 compares hearing thresholds for HI and NH participants at 0.25, 1, and 4 kHz frequencies among the three conditions with decreasing amount of supervision. In general, median thresholds of conditions II and III were in line with those of condition I for all groups and frequencies. As expected, median thresholds of HI were higher than NH for all three frequencies. Furthermore, median thresholds of HI listeners at 4 kHz were the highest, followed by 1 kHz and 0.25 kHz.

**Fig. 2.**
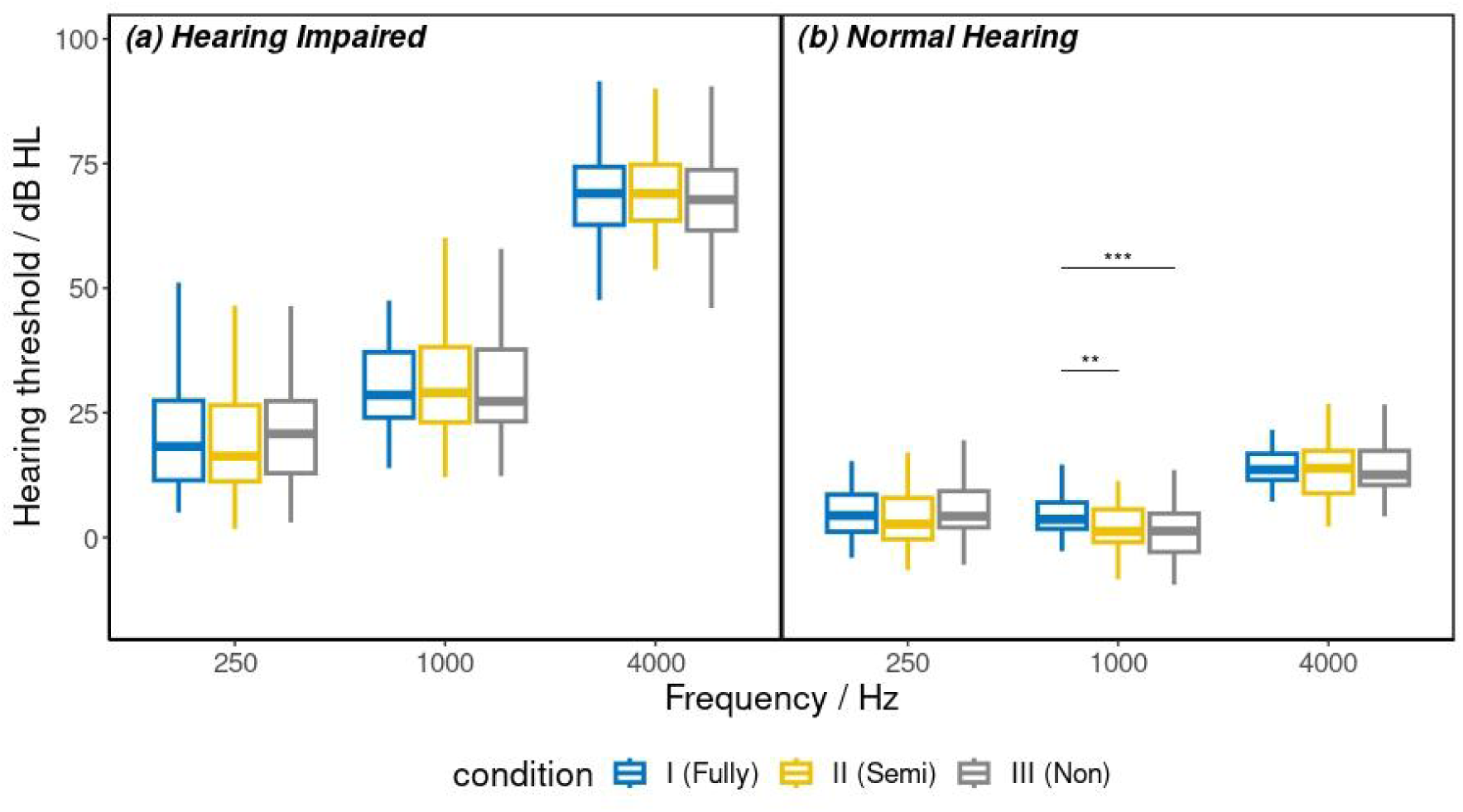
Hearing threshold L_50_ (in dB HL) grouped by three conditions (I: Fully-, II: Semi-, III: Non-supervised) for (a) hearing impaired (HI) and (b) normal hearing (NH) listeners as a function of three frequencies (0.25, 1, and 4 kHz). Condition I was set up as a reference. The medians, 25%, 75% percentiles, and interquartile ranges (IQR) are given in the respective bar-and-whiskers plot. The ends of the whiskers describe values within 1.5*IQR of the 25% and 75% percentiles. In case of statistically significant differences, the level of significance is labeled with stars above the lines.

A three-way mixed-design ANOVA was performed to analyze the effect of the degree of hearing loss (NH/HI), frequency (0.25, 1, and 4 kHz), and condition (I/II/III) on hearing threshold L_50_, revealing that there was a significant difference in hearing thresholds for the degree of hearing loss (p < 0.05) and frequency (p < 0.05) while no significant difference for condition (p = 0.22) was detected. The post-hoc analysis compared hearing thresholds of conditions II and III against I, indicating that conditions II and III did not significantly differ from condition I for all three frequencies within both listener groups except for the NH group at 1 kHz.

Statistical values, i.e., the signed difference, its absolute value, RMSE, R, and p value significance level of the thresholds L_50_ of conditions II and III against the reference condition I for two listener groups and three frequencies, are summarized in Table 3. Comparing the thresholds between conditions II and I (upper half of Table 3), mean signed differences were less than 2 dB in most cases, while mean absolute differences were around 3 dB. All RMSE values were smaller than 5 dB. The R values of HI listeners were higher than 0.9, suggesting a strong positive correlation while the R values of NH listeners were higher than 0.65, indicating a moderately positive correlation. Regarding the comparison of L_50_ between conditions III and I (bottom half of Table 3), mean signed differences were less than 1 dB except for NH listeners at 1 kHz. Similar to the comparison between conditions II and I, the mean absolute differences were around 3 dB, RMSE values were less than 5 dB and there was a strong correlation in the NH group while a moderately positive correlation was found in HI listeners.

**Table 3.**
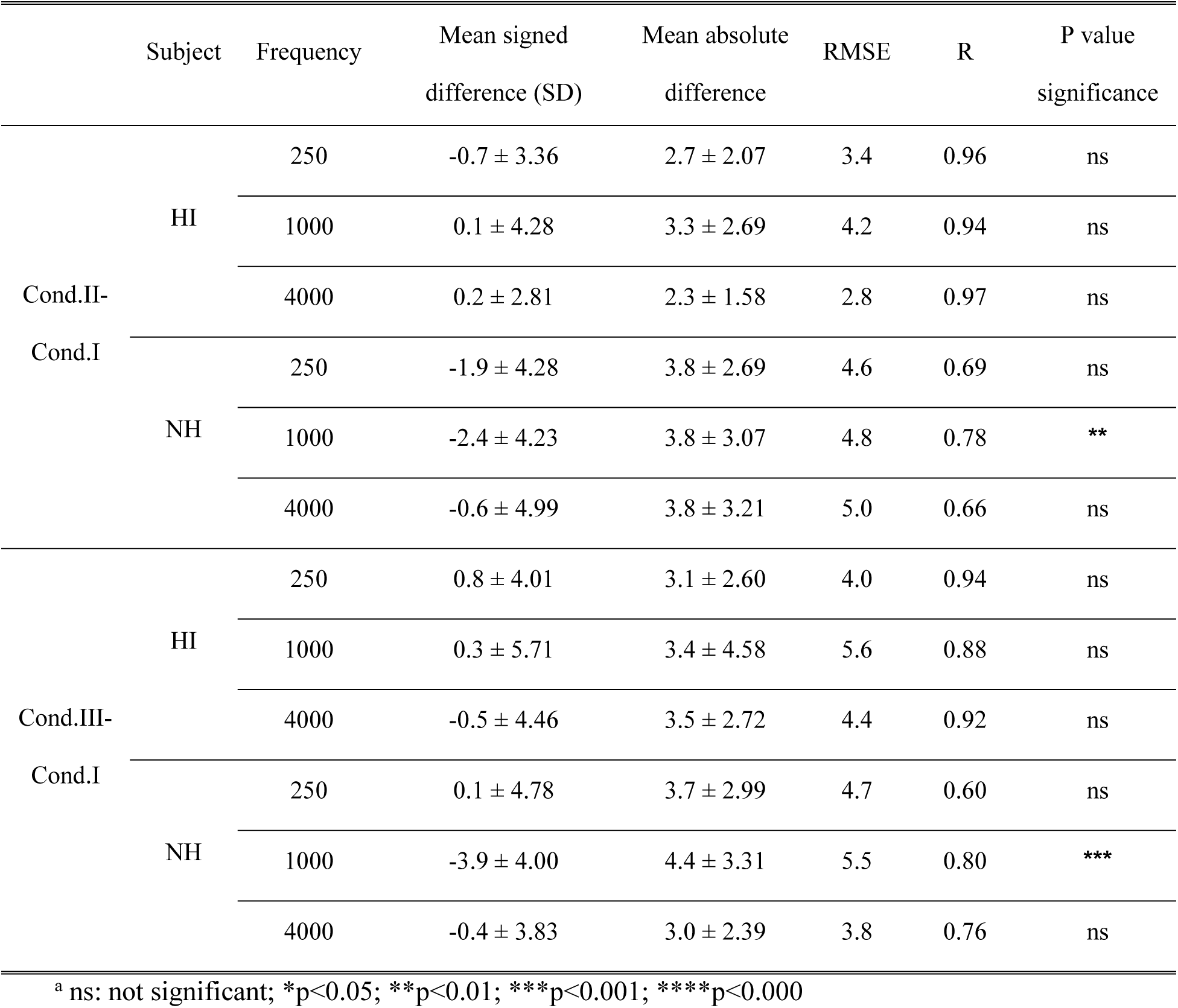
Quantitative comparison of the measured thresholds L_50_ between conditions II and I, and III and I in terms of signed difference, absolute difference, RMSE, R, and p value significance^a^ level.

### Experiment II: Adaptive Categorical Loudness Scaling

Fig. 3 plots the average loudness function of three conditions for HI and NH listeners at 0.25, 1, and 4 kHz frequencies. For all frequencies and listener groups, the average loudness functions of conditions II and III were consistent with condition I. The average loudness functions of HI listeners generally showed steeper growth than NH listeners, especially at 4 kHz, which could be explained by the ‘loudness recruitment’, as mentioned above. HI listeners exhibited a significant increase in the slope of the loudness function with an increase in frequency which was not observed in NH listeners.

**Fig. 3.**
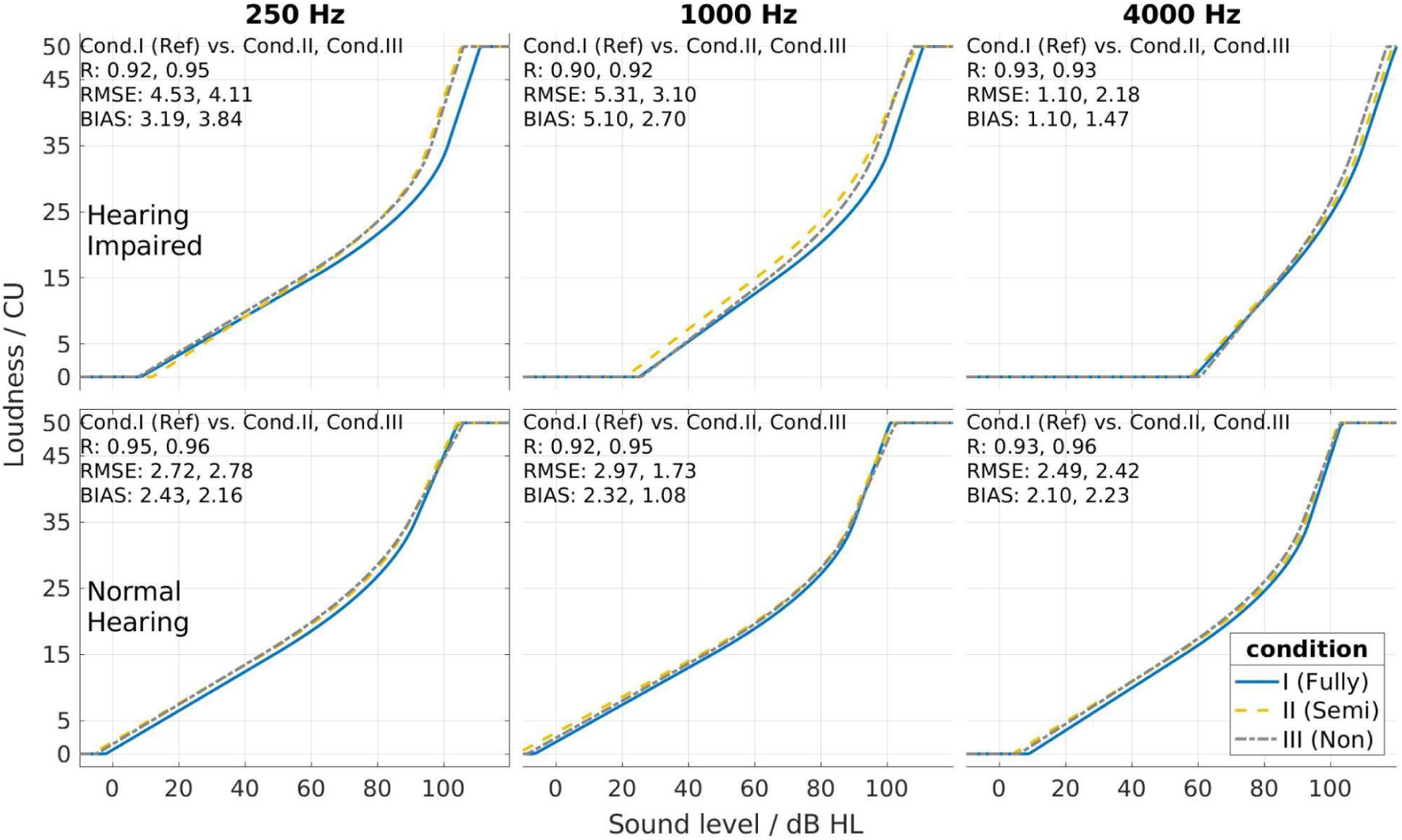
Average loudness growth function (i.e., loudness in CU as a function of sound level in dB HL) of the three experimental conditions employed (condition I = fully-supervised; II = semi-supervised; III = non-supervised) for HI (upper row) and NH (bottom row) listeners at 0.25 kHz (left column), 1 kHz (middle column), and 4 kHz (right column). The Pearson correlation coefficients (R), root mean square errors (RMSE), and biases between two conditions II and III against I (reference) of levels for each category units are provided in the upper left corner of each sub-figure.

Quantitatively speaking, the Rs of conditions II/III against I were higher than 0.9 for both NH and HI listeners at all three frequencies, indicating a rather high correlation of average loudness functions between conditions II and I, and between conditions III and I. HI listeners exhibited RMSE values less than 5 dB for most of the cases except for the comparison between conditions I and II at 1 kHz. NH listeners even produced a less than 3 dB RMSE value for all cases. Similarly, the bias for HI listeners was less than 4 dB and for NH listeners less than 3 dB with one exception occurring for HI listeners between conditions I and II at 1 kHz. Overall, the statistical measures suggested that the loudness function of conditions II and III showed a great agreement with condition I.

Five descriptive parameters (i.e., HTL, MCL, UCL, MLL, and DR) of three conditions for HI and NH listeners at 0.25, 1, and 4 kHz are shown in Fig. 4. The median descriptive parameters for all three frequencies and both listener groups in conditions II and III were close to the condition I. Moreover, the median levels of the five descriptive parameters did not change with an increase in frequency for NH listeners. As expected, the median levels of HTL increased while DR decreased with an increase in frequency for HI listeners. The IQRs of HTL and DR were larger for HI listeners compared to NH listeners.

**Fig. 4.**
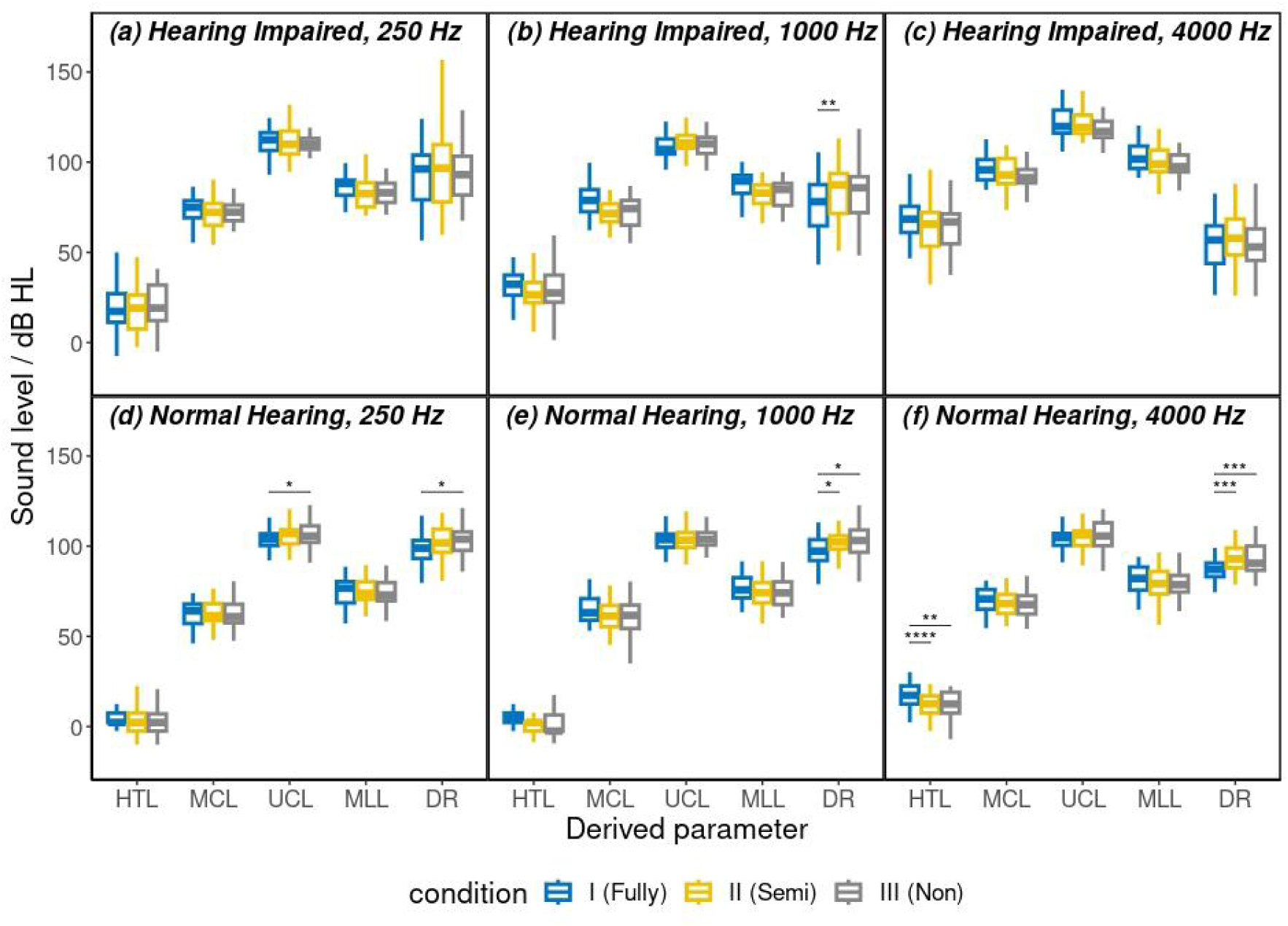
Five descriptive and intuitive parameters (in dB HL) derived from the loudness function of three conditions for HI and NH listeners at 0.25, 1, and 4 kHz frequencies. HTL: hearing threshold level (2.5 CU); MCL: most comfortable loudness level (20 CU); UCL: uncomfortable loudness level (50 CU); MLL: median loudness level (25 CU); DR: dynamic range (UCL-HTL). See Fig. 2 for an explanation of the bar-and-whiskers plot.

The effect of hearing impairment (NH/HI), frequency (0.25, 1, and 4 kHz), and condition (I/II/III) on five descriptive parameters (HTL, MCL, UCL, MLL, and DR) was assessed via the five different mixed-design ANOVA tests. The results revealed that there was a significant main effect of the degree of hearing loss and frequency on all five descriptive parameters (p < 0.05). Moreover, the factor condition was not significant on UCL (p = 0.12) while was significant on the other four descriptive parameters (p < 0.05).

A pair-wise t-test was performed to assess whether there was a significant difference in levels between conditions II and I, and III and I, respectively. For HI listeners, all five descriptive parameters of conditions II and III did not significantly differ from the condition I at all frequencies except for DR at 1 kHz. Furthermore, for NH listeners at 0.25 kHz, there was a significant difference in UCL and DR between conditions III and I (p < 0.05). At 1 kHz, the differences in DR between conditions II and I, and conditions III and I were significant. At 4 kHz, the differences across conditions of HTL and DR were significant.

As the t-test and the mixed-designed ANOVA test typically assume the ‘homogeneity of variance’ (i.e., all groups have the same variance), our data might violate the assumption (i.e., NH listeners have a smaller variance than HI listeners, as shown in Fig. 4), and thus the validity of the statistical tests might be affected. This would lead to falsely rejecting the null hypothesis (i.e., the factor condition is not supposed to be significant but reported to be significant).

Taken together, while for most cases the five parameters did not differ between the reference condition I and the less supervised conditions II and III, respectively, statistically significant differences only existed in a few groups, suggesting that these significant differences might not be systematic differences but rather random differences. In addition, the magnitudes of the significant differences in the NH and HI groups were overall less than 5 dB, indicating that the differences might not be clinically relevant. As we always measured condition I first, the sequence or training effect might explain such a difference.

### Experiment III: Binaural and Spectral Loudness Summation

#### Binaural loudness summation

Conditions I, II, and III are differentiated with three colors. Grey dashed line: 0 dB. LNN1000: one-third-octave-band centered at 1 kHz low-noise noise; UEN17: uniformly exciting noise at 17 critical bands.

Mean and standard deviation of the level differences for equal loudness (LDELs) as a function of loudness in CU of HI and NH participants for LNN1000 and UEN17 among three conditions are shown in Fig. 5. In most cases, the mean LDELs of conditions III and II were in agreement with those of condition I. It is notable that the standard deviation of LDEL of the condition III for LNN1000 at 25 and 50 CU for HI listeners was considerably larger than conditions II and I. Binaural loudness summation was signaled by mean LDELs significantly larger than 0, which was observed in most groups. Exceptions were observed for the HI listener at 2.5 and 50 CU of the condition III and NH listener at 2.5 CU of the condition II stimulated by LNN1000. Generally, the LDELs of 25 CU were the highest except for HI listeners of conditions II and III stimulated by UEN17.

**Fig. 5.**
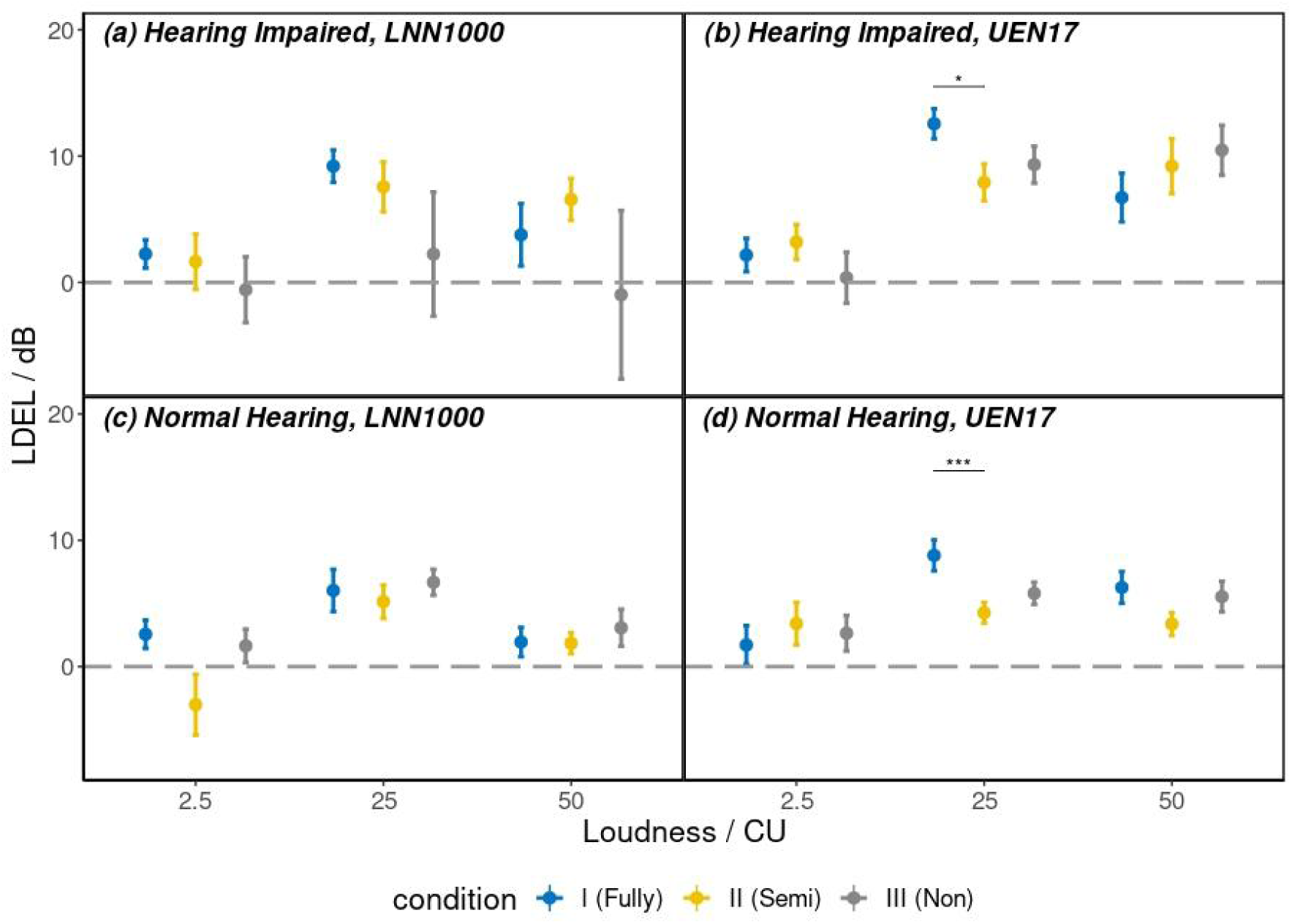
Mean and standard deviation (denoted by whiskers) of level difference for equal loudness (LDEL, in dB) between binaural and monaural (left ear) presentation for equal loudness at 2.5, 25, and 50 CU using narrowband noise (LNN1000) and UEN17 broadband noise, respectively, for HI (upper row) and NH (bottom row) listeners.

A four-way mixed-design ANOVA (NH/HI as a between-subject factor, condition (I/II/III), frequency (0.25, 1, and 4 kHz), and loudness (2.5, 25, and 50 CU) as within-subject factors) was conducted to assess the effect on LDEL. There was a significant main effect on the degree of hearing loss (p < 0.05), frequency (p < 0.05), and loudness CU (p < 0.05). The main effect of the condition was, however, not significant (p = 0.4).

Despite the insignificant main effect of the condition, the post-hoc analysis employing a pairwise t-test with ‘Bonferroni’ adjustment was carried out on LDEL, where condition I was the reference. In general, the LDEL of conditions II and III did not differ from condition I. However, a significant difference occurred in some pairs, i.e., between conditions I and II at 25 CU for both NH (p < 0.05) and HI (p < 0.001) stimulated by the UEN17 broadband signal. Even though these differences were statistically significant, the mean values of the differences were roughly 6 dB. Thus, similar to the results above, the significant differences in statistics might not be clinically relevant differences.

#### Spectral loudness summation

Fig. 6 shows LDEL (with error bars) of three conditions as a function of loudness in CU between LNN250 and UEN17 (left), LNN1000 and UEN17 (middle), and LNN4000 and UEN17 (right) for HI (upper) and NH (bottom) listeners. Generally, the mean difference of LDEL between conditions II and I, and between III and I was small with values smaller than 10 dB. For HI listeners, the mean LDELs at 25 and 50 CU were greater than 0 while lower than 0 at 2.5 CU concerning the comparison between LNN250 and UEN17. However, the mean LDELs of NH listeners were larger than 0 at three CU. Comparing the LDELs between LNN1000 and UEN17, both NH and HI listeners exhibited a negative LDEL at 2.5 CU while positive at 25 and 50 CU for three conditions with one exception of the HI listener for the condition III at 50 CU. Regarding the mean LDEL difference between LNN4000 and UEN17, NH and HI participants showed a substantial difference: the mean LDELs of HI listeners were always positive, while NH listeners were around 0.

**Fig. 6.**
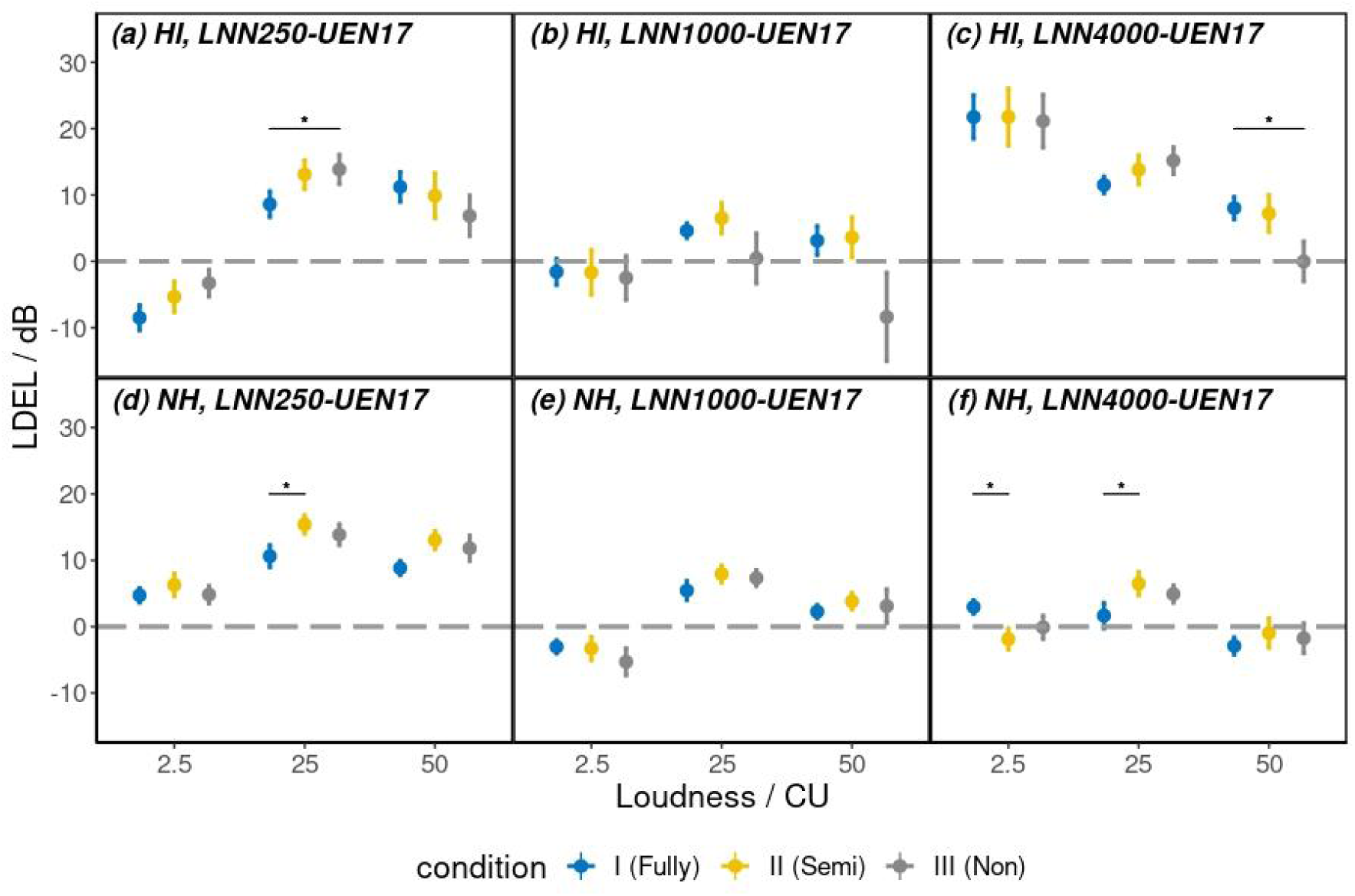
Mean and standard deviation (denoted by whiskers) of level difference for equal loudness (LDEL) between three narrowband stimuli (LNN250, left; LNN1000, middle; LNN4000, right) and one broadband stimulus (UEN17) for equal loudness at 2.5, 25, and 50 CU for HI (upper row) and NH (bottom row) listeners. Grey dashed line: 0 dB. All signals were presented monaurally on the left ear. LNN250, 1000, 4000: one-third-octave-band centered at 0.25, 1, and 4 kHz low-noise noise; UEN17: unified excitation noise at 17 critical bands.

A four-way mixed-design ANOVA was conducted to assess the effect of the degree of hearing loss (HI/NH), condition (I, II, and III), comparison (LNN250-UEN17, LNN1000-UEN17, LNN4000-UEN17), and loudness (2.5, 25, and 50 CU) on LDELs, in which the first factor was set up as a between-subject factor while the latter three as within-subject factors. The statistical outcome of ANOVA revealed that the main effect of all four factors was significant (p < 0.05).

A pair-wise t-test as a post-hoc analysis was performed to check whether the LDEL between conditions II/III and I was significantly different. The results suggested that in most cases, the LDEL of conditions II and III did not significantly differ from the LDEL of the condition I. For HI participants, there was only a significant difference between conditions I and III on LDEL at 25 CU in comparison pairs of LNN250-UEN17 and at 50 CU of LNN4000-UEN17 (p < 0.05). For NH listeners, only the difference in LDEL between conditions I and II was significant at 25 CU of LNN250-UEN17, and at 2.5 and 25 CU of LNN4000-UEN17 (p < 0.05).

## Discussion

Performing pure-tone audiometry and categorical loudness scaling on a smartphone was demonstrated here to be feasible if the smartphone is calibrated properly, the ambient noise is under control and the adaptive procedure provides high precision. The test outcome on a smartphone appears to be valid since it is aligned with the laboratory measurement in most cases. The way of supervision does not have a general impact on the measurement results, i.e., the non-supervised automated tests performed here are in principle equivalent to the fully-supervised manual tests.

The smartphone hearing tests employed here are applicable and accessible not only for normal hearing participants but also for persons with a hearing loss. It is useful and not difficult for HI listeners to administer the measurements themselves on a smartphone if they are familiar with the procedures. On top of the commonly employed unaided ACALOS measurement, i.e., narrowband signal presented unilaterally, the broadband stimulus for binaural presentation is also evaluated on a smartphone and does not show a large difference compared to the lab test. The usage of a variety of stimuli for adaptive categorical loudness scaling might support fine-tuning for a non-linear hearing aid on a smartphone in the future.

### Pure-tone audiometry

A number of studies have considered the difference between an app-based tone-in-quiet measurement and the clinical audiogram in order to validate the respective app on the mobile device. They either do the comparison in a clinical sound-insulated environment for both cases (e.g., Swanepoel et al., 2010; Colsman et al., 2020; Hazan et al., 2022) - which is supposed to yield no difference due to the acoustic presentation mode – or in a “quiet everyday environment” for the app (e.g., Kam et al., 2012; Abu-Ghanem et al., 2016) where any observed difference may be due to acoustical reasons (i.e., low-frequency noise components that can hardly be suppressed by ear-level devices), due to procedural differences (e.g., distraction due to attention-demanding occurrences in daily life, see Xu et al., 2023), or due to device calibrations. Typically, those studies that perform the validation under similar clinical, acoustically controlled, and distraction-sparse conditions as in our study agree with our study by reporting only a very small mean (signed) difference (e.g., within 5 dB as revealed in Thai-Van et al., 2022) across conditions. In addition, in a relatively noisy environment, fewer differences are expected for hearing-impaired listeners since their audiometric results would only be affected by higher ambient noise levels than normal-hearing listeners. As pointed out by Swanepoel et al. (2010), as HI listeners typically have a reduced hearing sensitivity, the apparent awareness of the internal noise level in NH listeners is largely eliminated.

### Adaptive categorical loudness scaling

To our knowledge, there is no study so far evaluating categorical loudness scaling on a smartphone. Our experimental results provide the first evidence that it is plausible and valid to perform non-supervised CLS measurement on a smartphone both for NH and HI listeners. In addition, there is only one study so far, i.e., Kopun et al. (2022), which evaluated the CLS measurement on a laptop remotely in comparison to a clinical database. This is comparable with the comparison between conditions II and I in our study on the group level. Kopun et al. (2022) reported that for NH participants (N = 5), the mean signed difference averaged across categories was 5.9 and 4.9 at 1 and 4 kHz, respectively. The mean signed difference of our study is much smaller, i.e., 2.3 and 2.1 for 1 and 4 kHz. First, the fitting of the loudness function might play a role. Kopun et al. (2022) simply calculated the median level of each category to describe the individual loudness function without fitting the data to a 2-segment linear function. Second, the outliers were not removed, leading to non-monotonic loudness growth. This contrasts to our study where we fitted the individual responses based on the method introduced in Oetting et al. (2014) to obtain an individual monotonic loudness function. Third, the test environment might make an impact. We conducted all experiments in a sound-attenuated booth to eliminate the influence of environmental noise. Kopun et al. (2022), however, did in-lab measurements at a sound-treated booth while remote laptop measurements at home. Although Kopun et al. (2022) attempted to control and check the noise level between runs in the remote measurements, the fluctuating environmental noise might influence the loudness judgment during the run. Fourth, Kopun et al. (2022) used a different calibrated headphone (i.e., Sennheiser HD 280 Pro). Lastly, the time gap between conditions II and I in Kopun et al. (2022) ranged from 2 years 6 months to 2 years 9 months while our time gap was less than a day. Overall, these differences not only in the experimental setup but also in the data processing would explain why our study exhibits a higher reproducibility than the earlier study, indicated by a smaller mean signed difference.

The descriptive parameters (i.e., HTL, MLL, UCL, and DR) of our study measured with a smartphone for NH listeners match quite well with the reference values reported in Oetting et al. (2016). The mean difference of the 4 parameters between Oetting et al. (2016) (N = 9) and our results is less than 2 dB at 0.25 kHz while lying within one standard deviation at 1 and 4 kHz. Furthermore, our measured MLLs and DRs are quite consistent with the empirical values for young NH listeners (N = 11) and HI listeners (N = 70) provided by Sanchez-Lopez et al. (2021). The median MCLs and DRs of NH listeners reported by Sanchez-Lopez et al. (2021) were 70 and 97.5 dB HL at low frequencies, and 75 and 92.5 dB HL at high frequencies while the median MCLs and DRs of listeners measured by us were 73.5 and 103.5 dB HL for low frequencies, and 78.7 and 90.6 dB HL for high frequencies. The difference between Sanchez-Lopez et al. (2021) and our study is around 5-6 dB and relatively small. Comparing the HI listeners of Sanchez-Lopez et al. (2021), most of our measured parameters for both low and high frequencies stay within the 25% and 75% percentile range of Sanchez-Lopez et al. (2021) except for MCLs at high frequencies. One possible reason might be different high frequency measurements: we only measured 4 kHz while Sanchez-Lopez et al. (2021) measured 2, 4, and 6 kHz and averaged the values of MCL. Another explanation could be that individual (within-subject) preference for MCLs might vary. Overall, the descriptive parameters measured by a smartphone show good consistency with the empirical values reported in the literature for both NH and HI listeners.

The three conditions differing in degree of supervision with calibrated hardware appear not to systematically influence the results of CLS in terms of both loudness growth functions and derived parameters (as shown in Fig. 3 and revealed by the mix-designed ANOVA), implying that we could let the participants test themselves on a smartphone for the CLS test, which meets our expectations. One reason to explain the results might be that the task for loudness judgment is rather intuitive and natural based on the feedback from our participants covering both NH and HI listeners. In addition, CLS is a supra-threshold measurement, which is expected to be less prone to influence by factors such as hardware and environment. Unlike some other speech-related tasks, e.g., the speech-in-noise test or listening effort test which are rather cognitively demanding, the CLS task does not involve speech comprehension, and, therefore, should be rather robust without any additional assistance from experimenters.

### Binaural and Spectral Loudness Summation

Level differences for equal loudness (LDELs,) - that quantify the binaural and spectral loudness summation - mostly do not show differences between the standard in-lab and smartphone measurements. This indicates that the smartphone measurements could detect the binaural and spectral loudness summation as well as the assessment conducted in a laboratory. However, we find that the factor condition shows a significant main effect on LDEL for the spectral loudness summation and in some groups, there is a significant difference in LDEL between conditions, as revealed by the post-hoc t-tests. Despite the (unexpected) significant difference in statistics, the values of the difference in LDEL are generally below 10 dB, which might not be considered to be clinically significant (e.g., in Thai-Van et al., 2022, 10 dB difference is defined as a criterion to determine the ‘clinical equivalence’).

A similar amount of binaural loudness summation for NH listeners can be observed in our study as reported by Oetting et al. (2016), indicating that the binaural LDELs for both broadband and narrowband signals are highest at 25 CU and lowest at

2.5 and 50 CU. Furthermore, the broadband signal exhibits higher LDELs than the narrowband signal. For broadband signals, a higher individual variability at high loudness could be observed for HI than for the NH listeners, which is compatible with Oetting et al. (2016). Whilby et al. (2006) examined 1-kHz pure tones for HI listeners, suggesting that LDELs were around 6 dB at medium loudness levels, decreased towards lower levels, and exhibited high individual variability. Their findings are quite comparable with our results, although we employ a different stimulus (i.e., 1 kHz one-third octave noise).

Concerning the spectral loudness summation experiment, our results in general are in line with Brand and Hohmann (2001). They reported that spectral LDELs were around 25 dB for speech shaped noise at medium loudness, and decreased towards lower and higher loudness for NH listeners (N = 8). We have a similar trend but smaller values of LDELs. This might be explained by the applied broadband signal: in our case, it is UEN17 while speech-shaped noise with different speech spectra was employed by Brand and Hohmann (2001). For HI listeners (N = 8), Brand and Hohmann (2001) showed that LDELs were approximately 10 dB and decreased with lower loudness, which is in line with our results.

Loudness scaling and loudness matching appear to be the two main tools to assess loudness summation for practical applications. Van Beurden et al. (2021) compared the two measurement procedures and concluded that both procedures provided valid and reliable results. Loudness scaling, on one hand, provides information on the entire loudness range. It requires a simple categorical judgment task, which is quite intuitive even for the elderly and naïve participants while loudness matching is less intuitive and needs more instructions for the listeners who have to “equalize apples and pears”, i.e., are forced to judge two differently perceived stimuli as being equal in one domain which is a challenge for inexperienced persons. On the other hand, loudness scaling might be more time-consuming than loudness matching. Even though we do not systematically compare the two methods on a smartphone, we prefer to apply loudness scaling on mobile devices since the feedback from our participants indicates that it is rather straightforward and easy to measure while using an acceptable measurement time.

### Limitations and outlook

One major limitation of the pure-tone audiometry in this study is that we only measured three frequencies, which mainly cover the speech range. For more refined clinical diagnostics, it might be of interest to measure in total 11 frequencies for both ears similar to the clinical audiogram. However, for a rough classification of hearing loss and given the limited additional information of additional audiogram frequencies at the cost of a higher time effort, the choice of three frequencies is a compromise.

Our current study only considers conducting the smartphone measurements in a sound-treated booth in order to eliminate any effects of the environment on the measurement outcome (e.g., distraction or background noise). It is worthwhile to consider experiments outside the booth while still ensuring the quality of the audiometric data. A possible solution could be monitoring the real-time noise level during the measurement as Kopun et al. (2022), Swanepoel et al. (2014; 2015), Maclennan-Smith et al. (2013), and Serpanos et al. (2022) did. Another approach for out-of-booth measurement could be using noise cancellation earphones (e.g., Clark et al., 2017).

The headphone employed here is a professional audiometric headphone (Sennheiser HDA200), which appears to be expensive and not publicly accessible. Van der Aerschot et al. (2016) recommended that affordable headphones, e.g., Sennheiser HD202 could be applied for pure-tone audiometry assessment. Moreover, the cheap headphones Sennheiser HD 280 Pro circumaural headphone was utilized by Kopun et al. (2022). Pickens et al. (2018) suggested that both, the Pioneer HDJ-2000 (Pioneer, Bunkyo, Tokyo, Japan) and the Sennheiser HD280 Pro (Sennheiser, Wedemark, Hanover, Germany) headphones, could be employed for mobile pure-tone audiometry assessment. The true wireless stereo (TWS) earbuds for pure-tone audiometry introduced by Guo et al. (2021) might also be considered as a daily-accessible alternative to the audiology headphone.

In our current study, we calibrated the smartphone output accurately in order to eliminate the influence of calibration and make it comparable to the standard laboratory measurement. However, in everyday life, the smartphone is normally not calibrated.

How to treat the uncalibrated mobile device and additional hardware in non-laboratory setups remains a challenge. Kisić et al. (2022), for instance, proposed that human speech might be an appropriate and stable test signal for microphone calibration while Scharf et al. (2023) considered the whistling sound of a 0.33 l beer bottle as a rough calibration signal.

## Conclusions

Three different experiments were designed to validate the usage of smartphone-based, non-supervised audiometric tests by studying the influence of the degree of supervision on audiometric tests to be performed with mobile devices:

- Experiment I (pure-tone audiometry) indicates that the way of supervision does not influence the measurement outcome. More specifically, the mean signed difference and mean absolute difference between smartphone and laboratory audiometry of NH and HI listeners exhibit less than 1 dB and 4 dB, respectively, in most cases.
- Experiment II (Adaptive CLS measurement) reveals that supervision does not affect the outcome values of categorical loudness scaling (i.e., the derived loudness growth functions of NH and HI listeners). The bias between smartphone and in-lab loudness function is considerably small and yields 2.67 and 1.8 dB for NH and HI participants, respectively. In addition, the 5 intuitive parameters (i.e., HTL, MCL, MLL, UCL, and DR) of smartphone CLS do not differ from the standard CLS assessment.
- Experiment III (binaural and spectral loudness summation) implies that binaural and spectral loudness summation can be derived by employing a smartphone in a way consistent with lab experiments. The LDELs measured on a smartphone between unilateral and bilateral presentation to quantify binaural loudness summation for both NH and HI listeners concerning both narrowband and broadband signals are consistent with those measured inside an acoustics laboratory. A similar trend is observed for the spectral loudness summation. Furthermore, the individual variations of HI listeners in loudness summation at loudness uncomfortable levels for binaural broadband signals are considerably large. Thus, in line with Oetting et al. (2016), including a binaural broadband signal for measuring the loudness perception appears to be a valid prerequisite for hearing aid fitting.

In conclusion, both audiometric tests considered here can be used for non-supervised smartphone-based hearing examination and are expected to yield very similar results as being conducted in a controlled laboratory experiment.

## Data Availability

All data produced in the present study are available upon reasonable request to the authors.

## Acknowledgments

This work was funded by the Deutsche Forschungsgemeinschaft (DFG, German Research Foundation) under Germany’s Excellence Strategy – EXC 2177/1 - Project ID 390895286.

